# Genetic and environmental effects on weight gain from young adulthood to old age and its association with body mass index at early young adulthood: an individual-based pooled analysis of 16 twin cohorts

**DOI:** 10.1101/2025.05.28.25328482

**Authors:** Alvaro Obeso, Gabin Drouard, Aline Jelenkovic, Juan R Ordoñana, Juan F Sánchez-Romera, Lucía Colodro-Conde, Miina Ollikainen, Sari Aaltonen, Robin P Corley, Brooke M Huibregtse, Emanuela Medda, Corrado Fagnani, Virgilia Toccaceli, Margaret Gatz, David A Butler, Meike Bartels, Lannie Ligthart, Eco JC de Geus, Kaare Christensen, Axel Skytthe, Kirsten O Kyvik, Sarah E Medland, Scott D Gordon, Finn E Rasmussen, Per Tynelius, Carol E Franz, William S Kremen, Michael J Lyons, Timothy Spector, Massimo Mangino, Genevieve Lachance, Patrik KE Magnusson, Nancy L Pedersen, Anna K Dahl Aslan, Glen E Duncan, Dedra Buchwald, Hyojin Pyun, Jooyeon Lee, Soo Ji Lee, Joohon Sung, Susanne Bruins, René Pool, Anders Eriksson, Nicholas G Martin, Dorret I Boomsma, Jaakko Kaprio, Karri Silventoinen

## Abstract

**Introduction:** Genetic factors contribute to weight gain, but how these effects change over adulthood is still unknown. We studied the impact of genetics on BMI change from young adulthood to old age and its relationship with BMI in early young adulthood.

**Data and Methods:** Data from 16 longitudinal twin cohorts, including 111,370 adults (56% women) and 55,657 complete twin pairs (42% monozygotic), were pooled. The data were divided into three stages (young adulthood-early middle age, late middle age, and old age). BMI change was calculated using linear mixed effects and delta slope methods. Genetic and environmental contributions to these changes and their correlations with baseline BMI were estimated through structural equation modeling.

**Results:** The average BMI increase per year was 0.18 kg/m² in men and 0.15 kg/m² in women during young adulthood-early middle age (18–50 years), decreasing to ≤0.07 kg/m² at older ages. Genetic effects contributed to BMI change during young adulthood-early middle age (men a²=0.29; women a²=0.26) and less so in late middle age (51–64 years) (men a²=0.05; women a²= 0.16) and old age (>65 years) (men a²=0.13; women a²= 0.18). Most variation was explained by non-shared environmental effects. In men, greater BMI during early young adulthood (18–30 years) was associated with lower BMI change later in life (r= –0.22 to –0.13), and the association was driven by genetic (r_A_=–0.27) and non-shared environmental (r_E_=–0.22 to –0.14) factors. In contrast, the association was positive in women (r=0.05–0.28) and was explained by genetic factors (r_A_=0.27– 0.51).

**Conclusion:** Genetics influence BMI change across adulthood, with their impact varying by age and sex. Environmental factors are the main drivers of adult BMI change, highlighting the role of modifiable factors in long-term weight regulation.

## Introduction

Body Mass Index (BMI) is the most widely used indicator of obesity in epidemiological studies. Moreover, high and increasing BMI across the lifespan increases health risks and is associated with increased mortality (1) and the risk of several diseases (1, 2, 3). While BMI has been extensively studied cross-sectionally, less is known about how genetic and environmental factors contribute to BMI change throughout adulthood. Longitudinal studies on BMI are essential to understand the processes underlying obesity risk in adulthood and can help when planning interventions to prevent weight gain. Studies on BMI trajectories suggest that BMI increases from early to mid-adulthood and either stabilizes or declines at later ages (4, 5). Overall, the trajectories across adult stages differ in men and women. These differences can be due to various factors, such as different exposures and life events, and different genetic factors affecting BMI in men and women as found in twin studies (6). Previous studies have reported differences in the genetics of weight gain trajectories during some stages of adulthood in twin cohorts (7, 8, 9). However, the influence of genetic and environmental factors on these patterns over adulthood is still poorly understood.

The influence of genetic factors on BMI and other obesity indicators is well-established (10, 11), and twin studies have estimated the heritability for BMI to vary from nearly 80% in early adulthood to approximately 60% in old age (6). Additionally, genome-wide association (GWA) studies have identified numerous genetic variants linked to BMI differences in adulthood based on cross-sectional analyses (12, 13). However, the genetic basis of weight gain remains unclear. Research involving twins and families has demonstrated that genetic factors contribute to weight change in adulthood, with heritability estimates ranging from 0.57 to 0.90 (9, 10). Studies based on Finnish twins have shown that the association between BMI in early adulthood and later BMI change is weak (8) or nonexistent in men and women (10). These and other previous studies have shown that genetic factors play a substantial role in baseline BMI (7, 10, 14) and BMI change (8, 9, 10, 14). However, the relative contributions of genetic and environmental influences on BMI change across the lifespan, how changes across different adulthood stages are linked to BMI in early adulthood, and the genetic and environmental correlations underlying these associations have not been thoroughly studied.

In this study based on pooled data from 16 longitudinal twin cohorts, we examined genetic and environmental influences on BMI change and how they differ from young adulthood-early middle age to older ages. Using the genetically informative twin design, we aimed to estimate i) differences across cohorts in how genetic and environmental effects explain individual differences in weight change, ii) how contributions of genetic and environmental effects differ between life stages from young adulthood to old age, and iii) how genetic and environmental factors contribute to the relationship between BMI in early young adulthood and subsequent weight changes.

## Data and methods

### Cohorts

The data were derived from the CODATwins database in collaboration with the BETTER4U consortium (15, 16). We selected 16 longitudinal twin cohorts from 10 countries across Europe, North America, East Asia, and Australia, all with BMI measures after 18 years of age. BMI was calculated as weight (kg) divided by height squared (m²). Descriptive information of the cohorts included in the study before applying exclusion criteria are displayed in Supplementary table 1.

Participants with at least two BMI measurements between 18 and 100 years were included, resulting in a pooled cohort of 111,370 twin individuals (56% women) from 55,657 complete twin pairs (42% monozygotic (MZ), 46% same-sex dizygotic (DZ), 12% opposite-sex DZ), with two to thirteen measurements. BMI was either measured (16%) or self-reported (84%). The Vietnam Era Twin Study of Aging had the smallest age range and fewest measures (15 years and 2 measurements), while the Danish Twin Cohort had the longest age range (82 years), and the Netherlands Twin Register had the most measurements (13 measurements). Cohort-specific data on baseline age, baseline BMI, and overall BMI change in men and women are presented in Supplementary Table 2, while Supplementary Table 3 provides cohort-specific BMI change information stratified by different stages of adulthood in men and women.

From the pooled data, different individuals were selected based on the analysis carried out. For the cohort-specific analyses, all individuals and measures were included. For the analyses stratified by life stage, individuals who had their first and last measurement in the same stage (young adulthood-early middle age: from 18 to 50 years old; late middle age: from 51 to 64 years old; old age: ≥65 years old) were included (N= 16, 219). For the study of phenotypic correlations and the genetic and environmental components underpinning these correlations, individuals who had their baseline measurement in what we defined as early young adulthood (from 18 to 30 years old) were included and then organized into the late middle adulthood and old age groups based on the age of their last BMI measurement (N= 10,171). Those who had their last measurement in young adulthood-early middle age were removed (N= 14,003).

Two sensitivity analyses were carried out. The first one, applied to all analyses, consisted of dividing the young adulthood-early middle age stage into three substages and old age into two stages, resulting in six stages: early young adulthood (from 18 to 30 years old), late young adulthood (from 31 to 40 years old), early middle age (from 41 to 50 years old), late middle age (from 51 to 64 years old), early old age (from 65 to 75 years old), and late old age (>75 years old), based on previously published literature (17, 18). The second sensitivity analysis focused on establishing associations, recognizing that changes in BMI are likely influenced not only by baseline BMI but also by other factors (such as zygosity, sex and baseline age among others). Beyond simply measuring the strength and direction of a linear relationship between two variables, this way of analyzing the relationship also accounts for potential causal or confounding influences, providing a broader perspective on the factors affecting BMI change.

### BMI change calculation

BMI change were calculated in two ways, depending on the number of measures of the individual. In both methods, an estimate of the rate of change in BMI (i.e., slope) and a baseline value (i.e., an intercept), collectively referred to as BMI trajectory, were obtained for each participant. The delta model was used for individuals with only two BMI measurements (in general or within a specific stage of life). This method subtracts the BMI values between two time points and then divides the differences in BMI by the time elapsed between them. The linear mixed-effects (LME) model was used for individuals with three or more longitudinal measures. LME models are suitable for analyzing longitudinal data as they include both fixed and random effects that account for correlations between longitudinal observations (19). In the LME models, individual trajectories were calculated using personal identifiers as random effects on the intercept and slope parameters. Outlier measures of BMI change were excluded (see the distribution of BMI change displayed in Supplementary figure 1). These analyses were performed using the R software (version 4.2.3) and the R packages lme4 (version 1.1-34), lmerTest (version 3.1-3), dplyr (version 1.1.4), modelsummary (version 1.4.3) and optimx (version 2023-10.21).

### Correlational analysis

Pairwise Pearson correlations were obtained to investigate the relationships between baseline BMI in early young adulthood (ages 18 – 30 years) and changes in BMI in later stages. The data from the included cohorts were pooled, and only twins with the first measurement in early young adulthood (aged 18 – 30 years) were included (N= 52,601 individuals). The data were then divided into three and six stages as detailed previously based on the age of the last measurement. Based on prior literature on sex differences (6, 8, 9) as well as evidence of significant sex differences in BMI trajectories found using LME models (results and the LME models are displayed in Supplementary Table 2), we stratified these analyses by sex. The non-random sample design was accounted for by using the family identifier as a random variable in the LME. Since changes in BMI are likely influenced not only by BMI at baseline but also by other factors, we conducted sensitivity analyses using LME model. In this model, the association between changes in BMI and BMI at baseline was examined with the addition of fixed-effect covariates. The model used is displayed in the caption of Supplementary Table 4 and 5. These analyses were performed using the R software (version 4.2.3) and the R package lme4 (version 1.1-34).

### Genetic structural equation modelling

Genetic twin modelling was performed to examine the genetic and environmental contributions to individual variations in BMI trajectories (20). MZ co-twins have virtually identical genetic DNA sequences, whereas DZ co-twins share, on average, 50% of their genes identical-by-descent. The trait variance can be decomposed into four components: (i) additive genetic effects (A), which include the effects of all the loci affecting the trait (correlation of 1 in MZ twins and 0.5 in DZ twins); (ii) genetic dominance effects (D), where the heterozygote phenotype deviates from the additive model; (iii) shared environmental effects (C), where all the environmental factors that make the co-twins similar are included (correlation of 1 in MZ twins and 1.0 in DZ twins); and (iv) unique environmental effects (E), in which all environmental variables making the co-twins different are included along with the measurement error (correlation of 0 in MZ and DZ twins). Univariate models were applied to identify the best fitting model for (i) BMI change across adulthood within each included cohort, (ii) BMI change during three stages in pooled data, and (iii) adulthood BMI change at six stages in pooled data. These models were also used to estimate the contributions of genetic and environmental effects. To identify the best-fitting model, intraclass twin correlations (ICC) for DZ and MZ co-twins were calculated by dividing within-pair variation by between-pair variation, as estimated through analysis of variance (Supplementary Tables 6 and 7). Based on these correlations, the additive genetic/shared environment/unique environment (ACE) model with both quantitative (different variance components for men and women) and qualitative genetic sex differences (the genetic correlation of opposite-sex DZ twins was freely estimated) was selected as the baseline model. This model was then compared to ACE models without sex differences, the ACE model without sex-specific genetic effects, and the AE model. The full AE model was used as a reference for comparison with AE models without sex differences and without sex-specific genetic effects. Model comparisons were conducted using −2 log likelihood (−2LL) tests and p-value information. Differences in model fit were assessed by comparing the χ^2^-distributed - 2LL measures for each model. Based on model comparisons, an AE model without qualitative sex difference but with different variance components for both sexes was selected for BMI change at (i) three stages and (ii) six stages (Supplementary Tables 8 and 9).

Finally, bivariate Cholesky decomposition, a model-free method to decompose all variations and covariations in the data into uncorrelated latent factors (21), was used to decompose the covariation between the three different measures (BMI at early young adulthood, BMI change from early young adulthood to late middle age and, BMI change from early young adulthood to old age) into genetic and environmental covariances. When these covariances were standardized, we obtained estimates of additive genetic (r_A_) and unique environmental (r_E_) correlations between BMI at early young adulthood and BMI change from early young adulthood to subsequent stages of adulthood. Genetic twin modeling was performed in the R software (version 4.2.3) and the R package OpenMx (version 2.21.11). The 95% confidence intervals (CI) were obtained by maximum likelihood estimation (22).

## Results

The mean baseline BMI (21.63 –28.88 kg/m^2^ in men and 21.69 –25.10 kg/m^2^ in women) varied between the cohorts. In most cases, mean baseline BMI values fell within the healthy weight range (BMI: from 18.5 to less than 25). The exceptions were the TwinsUK, Washington State Twin Registry, and Vietnam Era Twin Study of Aging cohorts in men, as well as both men and women from the Murcia Twin Registry. Across all cohorts, men had higher mean baseline BMI values compared to women (Supplementary table 2). The changes in BMI across the three adulthood stages by sex are presented in Table 1. In men and women, the mean rate of BMI change was higher during young adulthood-early middle age stage (0.18 kg/m^2^ per year for men and 0.15 kg/m^2^ per year for women) and then decreased being the lowest in old age (0.01 kg/m^2^ per year for men and - 0.01 kg/m^2^ per year for women). A similar decline in BMI change over aging was also observed when using six adulthood stages (Supplementary Table 10). Cohort-specific results showed substantial variation in BMI change among the cohorts in men (0.03 – 0.31 kg/m^2^ per year) and women (0.06 – 0.22 kg/m^2^ per year) (Supplementary table 2).

**Table 1.** BMI change rates (kg/m^2^ per year) across different stages of adulthood with 95% confidence intervals in the pooled data by sex. BMI change rate is displayed in young adulthood-early middle age, late middle age and old age stages in men and women from CODATwins data after the inclusion of the individuals with their first and last BMI measure both inside one of the three stages of adulthood. In both the change rate decrease as one progresses through the stages. **Abbreviations:** CI: Confidence Interval; LL: Lower limit; UL: Upper limit.

| Adulthood stages | Men |  |  | Women |  |  |
| --- | --- | --- | --- | --- | --- | --- |
|  | Mean | 95% CI |  | Mean | 95% CI |  |
|  |  | LL | UL |  | LL | UL |
| Young adulthood-early middle age (18-50 years old) | 0.18 | 0.16 | 0.24 | 0.15 | 0.11 | 0.21 |
| Late middle age (51-64 years old) | 0.04 | 0.01 | 0.07 | 0.06 | 0.02 | 0.10 |
| Old Age (>65 years old) | 0.01 | -0.01 | 0.03 | -0.01 | -0.05 | 0.04 |

The results of the relative proportions of additive genetic and unique environmental effects on BMI change in different cohorts are shown in Table 2. Additive genetic effects on BMI change were observed in men (a^2^=0.06 – 0.59) and women (a^2^=0.10 – 0.57) in all cohorts. However, for most of the cohorts, the unique environmental effects were even larger (e^2^ =0.41 – 0.94 in men and 0.43 – 0.93 in women). Moreover, we found significant heterogeneity in the additive genetic effects estimates across the cohorts (p < 0.001).

**Table 2.** Relative proportions of BMI change rate variance explained by additive genetic and unique environmental variance components with 95% confidence intervals of BMI change over the course of adulthood by cohort and sex. Full AE model was used in all the cohorts for BMI change variable **Abbreviations:** a^2^: Heritability; e^2^: proportion of total variance explained by unique environmental factors; CI: Confidence interval; LL: Lower limit; UL: Upper limit; BMI: Body mass index.

|  | Men |  |  |  |  |  | Women |  |  |  |  |  |
| --- | --- | --- | --- | --- | --- | --- | --- | --- | --- | --- | --- | --- |
|  | Additive genetics |  |  | Unique environment |  |  | Additive genetics |  |  | Unique environment |  |  |
|  | a<sup>2</sup> | 95% CI |  | e<sup>2</sup> | 95% CI |  | a<sup>2</sup> | 95% CI |  | e<sup>2</sup> | 95% CI |  |
|  |  | LL | UL |  | LL | UL |  | LL | UL |  | LL | UL |
| **Europe** |  |  |  |  |  |  |  |  |  |  |  |  |
| Danish Twin Cohort | 0.21 | 0.16 | 0.25 | 0.79 | 0.74 | 0.83 | 0.26 | 0.20 | 0.30 | 0.74 | 0.69 | 0.79 |
| Swedish Twin Register | 0.16 | 0.12 | 0.19 | 0.84 | 0.80 | 0.87 | 0.28 | 0.25 | 0.31 | 0.72 | 0.68 | 0.74 |
| Finnish Old Cohort | 0.21 | 0.16 | 0.25 | 0.79 | 0.75 | 0.83 | 0.25 | 0.20 | 0.28 | 0.75 | 0.71 | 0.79 |
| FinnTwin12 | 0.33 | 0.09 | 0.51 | 0.67 | 0.48 | 0.90 | 0.57 | 0.43 | 0.68 | 0.43 | 0.31 | 0.56 |
| FinnTwin16 | 0.47 | 0.37 | 0.54 | 0.53 | 0.45 | 0.62 | 0.56 | 0.49 | 0.62 | 0.44 | 0.37 | 0.50 |
| TwinsUK | 0.29 | 0.17 | 0.40 | 0.71 | 0.59 | 0.82 | 0.10 | 0.05 | 0.14 | 0.90 | 0.85 | 0.95 |
| Netherlands Twin Registry | 0.45 | 0.37 | 0.51 | 0.55 | 0.48 | 0.62 | 0.20 | 0.13 | 0.31 | 0.85 | 0.79 | 0.94 |
| Italian Twin Registry | 0.59 | 0.42 | 0.71 | 0.41 | 0.28 | 0.57 | 0.31 | 0.18 | 0.41 | 0.69 | 0.58 | 0.81 |
| Murcia Twin Registry | 0.15 | 0.00 | 0.33 | 0.85 | 0.66 | 1.00 | 0.07 | 0.00 | 0.16 | 0.93 | 0.88 | 1.00 |
| Swedish Young Male Twin Study | 0.51 | 0.45 | 0.55 | 0.49 | 0.44 | 0.54 | - | - | - | - | - | - |
| **North America** |  |  |  |  |  |  |  |  |  |  |  |  |
| Washington State Twin Registry | 0.24 | 0.18 | 0.31 | 0.76 | 0.68 | 0.81 | 0.22 | 0.16 | 0.28 | 0.78 | 0.71 | 0.83 |
| Colorado Twin Registry | 0.33 | 0.20 | 0.43 | 0.67 | 0.56 | 0.79 | 0.54 | 0.43 | 0.62 | 0.46 | 0.37 | 0.56 |
| NAS-NRC Twin Registry | 0.44 | 0.39 | 0.47 | 0.56 | 0.52 | 0.60 | - | - | - | - | - | - |
| Vietnam Era Twin Study of Aging | 0.06 | 0.00 | 0.16 | 0.94 | 0.83 | 1.00 | - | - | - | - | - | - |
| **Asia** |  |  |  |  |  |  |  |  |  |  |  |  |
| Korea Twin-Family Register | 0.20 | 0.11 | 0.29 | 0.80 | 0.74 | 0.87 | 0.10 | 0.00 | 0.21 | 0.90 | 0.78 | 1.00 |
| **Australia** |  |  |  |  |  |  |  |  |  |  |  |  |
| Queensland Twin Register | 0.21 | 0.12 | 0.28 | 0.79 | 0.71 | 0.87 | 0.16 | 0.10 | 0.20 | 0.84 | 0.79 | 0.89 |
| **World** |  |  |  |  |  |  |  |  |  |  |  |  |
| All cohorts | 0.31 | 0.29 | 0.32 | 0.69 | 0.67 | 0.70 | 0.19 | 0.17 | 0.20 | 0.81 | 0.79 | 0.82 |

Next, we studied the genetic and environmental contributions in the pooled data stratified by three adulthood stages (Table 3). We found evidence for additive genetic effects in all stages (a^2^= 0.05 – 0.29 in men and a^2^= 0.16 – 0.26 in women), with additive genetic effects being the highest in the young adulthood-early middle age stage. Additive genetic effects had the lowest values during late middle age in both sexes (a^2^= 0.05 and 0.16 in men and women, respectively), while in old age, the values were intermediate between the two previous stages (a^2^= 0.13 in men and 0.18 in women). The additive genetic effects were higher in women than in men, except in the young adulthood-early middle age. Similar results were found when considering six adulthood stages (Supplementary Table 11).

**Table 3.** Relative proportions of body mass index (BMI) changes (kg/m^2^ per year) rate variance explained by additive genetic and unique environmental variance components with 95% confidence intervals of BMI change across three stages of adulthood in the pooled data by sex. AE model without sex specific genetic effect was selected for BMI change in all the different stages of adulthood. **Abbreviations:** a^2^: proportion of total variance explained by additive genetic factors (Heritability); e^2^: proportion of total variance explained by unique environmental factors; CI: Confidence interval; LL: Lower limit; UL: Upper limit.

|  | Men |  |  |  |  |  | Women |  |  |  |  |  |
| --- | --- | --- | --- | --- | --- | --- | --- | --- | --- | --- | --- | --- |
|  | Additive genetic effect |  |  | Unique environmental effect |  |  | Additive genetic effect |  |  | Unique environmental effect |  |  |
| | $a^2$ | 95%CI | | $e^2$ | 95%CI | | $a^2$ | 95%CI | | $e^2$ | 95%CI | |
|  |  | LL | UL |  | LL | UL |  | LL | UL |  | LL | UL |
| Young adulthood-early middle age | 0.29 | 0.25 | 0.32 | 0.71 | 0.67 | 0.74 | 0.26 | 0.22 | 0.28 | 0.74 | 0.71 | 0.77 |
| Late middle age | 0.05 | 0.00 | 0.11 | 0.95 | 0.89 | 1.00 | 0.16 | 0.09 | 0.21 | 0.84 | 0.78 | 0.90 |
| Old age | 0.13 | 0.00 | 0.28 | 0.87 | 0.72 | 1.00 | 0.18 | 0.07 | 0.28 | 0.82 | 0.71 | 0.92 |

Finally, we analyzed whether BMI in early young adulthood (18 – 30 years old) is associated with later BMI change. Table 4 displays the phenotypic, additive genetic (r_A_), and unique environmental (r_E_) correlations. In men, a negative phenotypic correlation was observed between BMI in early young adulthood and BMI change from early young adulthood to subsequent stages (r= −0.22 – - 0.13), whereas in women, the correlations were all positive (r= 0.05 – 0.28). For the correlations between BMI in early young adulthood and BMI change from early young adulthood to subsequent stages, genetic factors contributed only in old age (r_A_= −0.27) and unique environmental factors in both late middle age (from 51 to 64 years old) and old age (≥65 years old) (r_E_= −0.22 – −014) in men. In women, only genetic factors in late middle age and old age accounted for the correlations (r_A_= 0.27 – 0.51). When these analyses were carried out using six adulthood stages, the phenotypic correlations showed the same directions except in men for BMI change from early young adulthood to early middle age (Supplementary table 12). The unique environmental factors showed similar results, except in women for BMI change from early young adulthood to late young adulthood. The additive genetic factors also showed similar results, being significant and positive in women in all stages, while in men, these factors did not reach statistical significance until early and late old age and presented a negative estimate. Sensitivity analysis, using LME models instead of Pearson correlations for exploring the relationships between BMI in early young adulthood and changes in the two subsequent stages of adulthood (late middle age and old age) (Supplementary table 4) and five subsequent stages of adulthood (Supplementary table 5) showed significant results in all cases except for women in late young adulthood. This reinforces the results obtained through Pearson correlation and enhances the robustness of our results, suggesting that the observed relationships persist even when accounting for potential confounding factors.

**Table 4.**
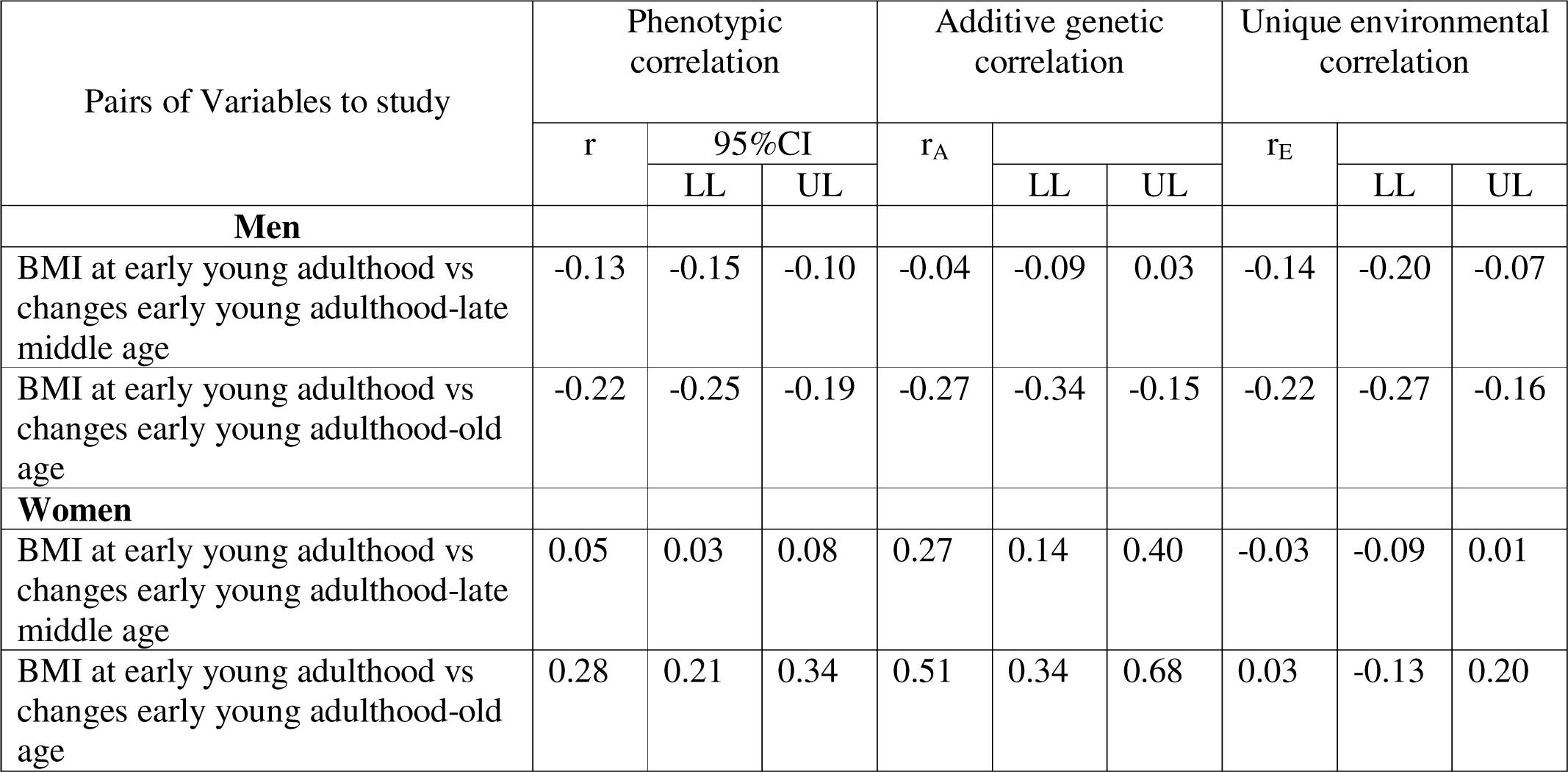
Phenotypic, additive genetic and unique environmental correlations of body mass index (BMI) at early young adulthood (18-30 years of age) and the changes in BMI (kg/m^2^ per year) from early young adulthood to subsequent stage in two stages of adulthood in the pooled data by sex. Pairwise correlations between body mass index (BMI) at early young adulthood and the changes in BMI (kg/m^2^ per year) from early young adulthood to subsequent stage in two stages of adulthood were carried out. They are summarized with correlations estimates besides their confidence intervals. Abbreviations: r_A_: additive genetic correlation; r_E_; specific environmental correlation; CI: Confidence interval; LL: lower limit; UL: upper limit; BMI: body mass index.

| Pairs of Variables to study | Phenotypic correlation |  |  | Additive genetic correlation |  |  | Unique environmental correlation |  |  |
| --- | --- | --- | --- | --- | --- | --- | --- | --- | --- |
| | r | 95%CI | | $r_A$ | | | $r_E$ | | |
|  |  | LL | UL |  | LL | UL |  | LL | UL |
| **Men** |  |  |  |  |  |  |  |  |  |
| BMI at early young adulthood vs changes early young adulthood-late middle age | -0.13 | -0.15 | -0.10 | -0.04 | -0.09 | 0.03 | -0.14 | -0.20 | -0.07 |
| BMI at early young adulthood vs changes early young adulthood-old age | -0.22 | -0.25 | -0.19 | -0.27 | -0.34 | -0.15 | -0.22 | -0.27 | -0.16 |
| **Women** |  |  |  |  |  |  |  |  |  |
| BMI at early young adulthood vs changes early young adulthood-late middle age | 0.05 | 0.03 | 0.08 | 0.27 | 0.14 | 0.40 | -0.03 | -0.09 | 0.01 |
| BMI at early young adulthood vs changes early young adulthood-old age | 0.28 | 0.21 | 0.34 | 0.51 | 0.34 | 0.68 | 0.03 | -0.13 | 0.20 |

## Discussion

This study examined the genetic and environmental contributions to BMI change over adulthood in multiple twin cohorts. The findings indicate that genetic effects play a role in BMI change; however, their influence varies across different stages of adulthood and differs between men and women. Genetic effects on BMI change were relatively higher in women, especially in the young adulthood-early middle age stage, while men showed lower genetic effects overall. Unique environmental effects emerged as the predominant influence across most cohorts. These findings highlight that while there is a genetic predisposition to BMI change, environmental effects linked to factors such as lifestyle, physical activity, medical conditions, and socioeconomic status, may exert a stronger and more sustained impact, especially in later adult stages. Furthermore, correlations observed between BMI in early young adulthood and BMI change in subsequent stages suggest long-term effects of early adult life factors on BMI regulation. In women, these correlations were positive while in men the correlations were negative. These results reinforce previous findings that early BMI is a predictor of later obesity and weight fluctuations (9).

Additive genetic factors influenced BMI change in all cohorts, although the proportion of variation explained by genetic effects varied among the cohorts. These differences could be due to factors related to the macro-environment, such as differences in demography, culture, eating and physical activity habits, in addition to variables like birth cohort and the age when measurements were taken. To the best of our knowledge, although studies on the heritability of BMI (12) and BMI change (8, 10, 14, 23) at different stages of adulthood have been conducted, no previous studies have reported the heritability of BMI change during adulthood in such large numbers of cohorts and life stages. Furthermore, after pooling the data and subsequent analysis of heritability in the different stages of adulthood in both men and women, the genetic effects exhibited two peaks of greater influence in both sexes: one in young adulthood-early middle age and the other in old age. Additionally, the lowest influence was observed in late middle age when studying three adulthood stages and early and late middle age when studying six stages. Previous twin studies have shown greater genetic effects on BMI (heritability ranged 0.57 – 0.77 in men and 0.59 – 0.75 in women) (6) than we found for weight change. This suggests that, while inherited traits contribute substantially to BMI, factors that include environmental effects (possibly individual life situations, lifestyle, marital status, working situation, and childbearing among others) are the primary drivers of BMI change throughout adulthood. Larger genetic effects on BMI change than in our study have been found in previous twin studies carried out in Finnish Twin cohorts (8, 10, 14) and other such as NAS-NRC Twin Registry (24) and Kaiser-Permanente Women Twins Study (California) (25). However, consistent results to those obtained in the current study were found in a previous study carried out in Danish and Chinese adult twins (including a total of 683 twin pairs) (23). Nevertheless, our study presents a major difference with the previously mentioned studies (the majority of them used a cohort included in the current study) which is the calculation of BMI change using a different method (delta method instead of LME or latent growth models).

Phenotypic correlations were identified between BMI in early young adulthood and BMI change from early young adulthood to subsequent stages in both men and women, but the direction of these correlations was different. The correlation coefficients for men were generally negative, indicating that those with higher BMI in early young adulthood tended to experience a decline in BMI change over time. Conversely, positive correlations were observed in women, indicating that those with higher BMI in early young adulthood tended to experience greater BMI change across subsequent stages of adulthood. Previously published twin studies that used some of the cohorts included in the currently used database have shown that correlations between BMI in early adulthood and its changes later in adulthood were weak (8) or non-existent (10).

Genetic and environmental factors have different contributions to the correlations between BMI in early young adulthood and later weight change in men and women. In men, negative genetic correlations were found between BMI in early young adulthood and its changes from early young adulthood to late middle age and old age. Moreover, significant negative unique environmental correlations were found in men between BMI in early young adulthood and changes from early young adulthood to the subsequent adulthood stages. These associations may arise if BMI in men in early young adulthood is primarily muscle mass, but weight gain is primarily fat mass accumulation especially considering that this life stage often coincides with peak physical fitness and higher engagement in physical activity. As men age, physical activity levels typically decline, and metabolic changes can lead to the development of sarcopenic obesity which is characterized by the increase in fat mass and a concurrent decrease in muscle mass (26). Therefore, the negative genetic and environmental correlations observed may indicate that individuals with higher muscle-related BMI in early young adulthood are genetically and environmentally predisposed to a lower rate of fat accumulation later in life. In women, all phenotypic correlations showed a positive significant genetic correlation, indicating consistent and substantial genetic overlap between BMI in early young adulthood and BMI change from early young adulthood to all adulthood stages. Significant environmental correlations were only observed when adulthood was divided into six stages specifically in BMI change from early young adulthood to early middle age. This suggests that unique environmental factors have a significant, stage-specific influence on BMI change.

The most important strength of this study is the extensive sample of twins with follow-ups from early young adulthood to old age, spanning over six decades with up to 13 measurements per individual. This allowed us to estimate the contributions of genetic and environmental factors on BMI change in different life stages as well as phenotypic, genetic and environmental correlations between BMI in early young adulthood and later changes in BMI until old age. Although we have used pooled data from most twin cohorts worldwide, there is a lack of data from many parts of the world, such as Africa and South America, and all cohorts represent high-income countries. Therefore, our results may only be generalized to affluent populations mainly following a Westernized lifestyle. Another limitation is the use of linear models to estimate changes in BMI during adulthood. BMI trajectories, as shown in the current study, may exhibit nonlinear patterns after 60 years old. Thus, the assumption that changes in BMI are linear over time can only be applied when the changes are observed between 18 and 60 years old, posing a limitation when including changes after the age of 60 years. Furthermore, we did not have information on modifiers, such as nutrition, physical activity, childbirth and other lifestyle factors that can contribute to BMI change. Finally, another limitation of the present study is related with the use of a stage ranging from 18 to 50 years of age. This broad age range covers several distinct periods of adult life, each characterized by different biological, psychological, and social processes, which may affect the interpretation of the BMI change patterns observed.

In conclusion, this study highlights the complex interplay between genetic and environmental effects on BMI change across adulthood in different cohorts and at various stages of adulthood. It shows that BMI in early young adulthood is linked to later changes, influenced by both genetic and environmental effects. Notably, genetic factors contribute less to variation in adult weight gain than previously suggested for BMI, and these genetic effects may differ from those affecting BMI in early young adulthood. The study also underscores important sex differences, emphasizing the need to consider sex-specific effects when examining BMI trajectories.

## Ethics statement

The pooled analysis was approved by the ethical board of the Department of Public Health, University of Helsinki. The data collections procedures of participating twin cohorts were approved by local ethical boards following the regulation in each country. Only anonymized data were delivered to the data management center at University of Helsinki.

## Data availability

The data used in this study is owned by the third parties (the individual twin cohorts) and made available to this study in condition that they will be used only in this meta-analysis. The data can be used based on the same principles as used in this study (more information from Karri Silventoinen).

## Code availability

All R scripts are available from the corresponding author.

## Author contributions

AO, GD, JK, and KS designed the study. JO, JSR, LCC, MO, SA, RC, BH, EM, CF, VT, MG, DB, MB, LL, EG, KC, AS, KK, SM, SG, FR, PT, CF, WK, ML, TS, MM, GL, PM, NP, ADA, GED, DB, HP, JL, SJL, JS, SB, RP, AE, NM, DIB, JK collected the original data files. KS, JK, AJ and AO pooled the data together. AO conducted the analyses and drafted the manuscript. GD, AJ, JO, JSR, LCC, MO, SA, RC, BH, EM, CF, VT, MG, DB, MB, LL, EG, KC, AS, KK, SM, SG, FR, PT, CF, WK, ML, TS, MM, GL, PM, NP, ADA, GED, DB, HP, JL, SJL, JS, SB, RP, AE, NM, DIB, JK and KS revised the manuscript. All authors approved the final version and agreed to be accountable for all aspects of the work in ensuring that questions related to the accuracy or integrity of any part of the work are appropriately investigated and resolved.

## References

1. Global Burden of Disease Study 2015. (2016). Global, regional, and national incidence, prevalence, and years lived with disability for 310 diseases and injuries, 1990–2015: A systematic analysis for the Global Burden of Disease Study 2015. Lancet, 388(10053), 1545–1602.

2. Wilborn, C., Beckham, J., Campbell, B., Harvey, T., Galbreath, M., La Bounty, P., Nassar, E., Wismann, J., & Kreider, R. (2005). Obesity: prevalence, theories, medical consequences, management, and research directions. Journal of the International Society of Sports Nutrition, 2(2), 4–31. 10.1186/1550-2783-2-2-4

3. Amin, V., Flores, C. A., & Flores-Lagunes, A. (2020). The impact of BMI on mental health: Further evidence from genetic markers. Economics and human biology, 38, 100895. 10.1016/j.ehb.2020.100895

4. Yang, Y. C., Walsh, C. E., Johnson, M. P., Belsky, D. W., Reason, M., Curran, P.,… & Harris, K. M. (2021). Life-course trajectories of body mass index from adolescence to old age: Racial and educational disparities. Proceedings of the National Academy of Sciences, 118(17), e2020167118.

5. Dahl, A. K., Reynolds, C. A., Fall, T., Magnusson, P. K., & Pedersen, N. L. (2014).Multifactorial analysis of changes in body mass index across the adult life course: a study with 65 years of follow-up. International journal of obesity (2005), 38(8), 1133–1141. 10.1038/ijo.2013.204

6. Silventoinen, K., Jelenkovic, A., Sund, R., Yokoyama, Y., Hur, Y. M., Cozen, W., Hwang, A. E., Mack, T. M., Honda, C., Inui, F., Iwatani, Y., Watanabe, M., Tomizawa, R., Pietiläinen, K. H., Rissanen, A., Siribaddana, S. H., Hotopf, M., Sumathipala, A., Rijsdijk, F., Tan, Q.,… Kaprio, J. (2017). Differences in genetic and environmental variation in adult BMI by sex, age, time period, and region: an individual-based pooled analysis of 40 twin cohorts. The American journal of clinical nutrition, 106(2), 457–466. 10.3945/ajcn.117.153643

7. Franz, C. E., Grant, M. D., Jacobson, K. C., Kremen, W. S., Eisen, S. A., Xian, H., Romeis, J., Thompson-Brenner, H., & Lyons, M. J. (2007). Genetics of body mass stability and risk for chronic disease: A 28-year longitudinal study. Twin Research and Human Genetics, 10, 537–545. 10.1375/twin.10.4.537

8. Drouard, G., Silventoinen, K., Latvala, A., & Kaprio, J. (2023). Genetic and Environmental Factors Underlying Parallel Changes in Body Mass Index and Alcohol Consumption: A 36-Year Longitudinal Study of Adult Twins. Obesity Facts, 16(3), 224–236. https://www.proquest.com/scholarly-journals/genetic-environmental-factors-underlying-parallel/docview/2900328328/se-2

9. Obeso, A., Drouard, G., Jelenkovic, A., Aaltonen, S., Palviainen, T., Salvatore, J. E., Dick, D. M., Kaprio, J., & Silventoinen, K. (2024). Genetic contributions to body mass index over adolescence and its associations with adult weight gain: a 25-year follow-up study of Finnish twins. International journal of obesity (2005). 10.1038/s41366-024-01684-3

10. Hjelmborg, J. V., Fagnani, C., Silventoinen, K., McGue, M., Korkeila, M., Christensen, K.,… & Kaprio, J. (2008). Genetic influences on growth traits of BMI: A longitudinal study of adult twins. Obesity, 16(4), 847–852.

11. Silventoinen, K., & Konttinen, H. (2020). Obesity and eating behavior from the perspective of twin and genetic research. Neuroscience & biobehavioral reviews, 109, 150–165.

12. Yengo, L., Sidorenko, J., Kemper, K. E., Zheng, Z., Wood, A. R., Weedon, M. N.,… & Frayling, T. M. (2018). Meta-analysis of genome-wide association studies for height and body mass index in ∼700,000 individuals of European ancestry. Human Molecular Genetics, 27(20), 3641–3649.

13. Wainschtein, P., Jain, D., Zheng, Z., TOPMed Anthropometry Working Group, NHLBI Trans-Omics for Precision Medicine (TOPMed) Consortium, Cupples, L. A., Shadyab, A. H., McKnight, B., Shoemaker, B. M., Mitchell, B. D., Psaty, B. M., Kooperberg, C., Liu, C. T., Albert, C. M., Roden, D., Chasman, D. I., Darbar, D., Lloyd-Jones, D. M., Arnett, D. K., Regan, E. A.,… Visschero, P. M. (2022). Assessing the contribution of rare variants to complex trait heritability from whole-genome sequence data. Nature genetics, 54(3), 263–273. 10.1038/s41588-021-00997-7

14. Ortega-Alonso A, Sipilä S, Kujala UM, Kaprio J, Rantanen T. Genetic influences on change in BMI from middle to old age: a 29-year follow-up study of twin sisters. Behav Genet. 2009;39(2):154–64.

15. Silventoinen, K., Jelenkovic, A., Sund, R., Honda, C., Aaltonen, S., Yokoyama, Y., Tarnoki, A. D., Tarnoki, D. L., Ning, F., Ji, F., Pang, Z., Ordoñana, J. R., Sánchez-Romera, J. F., Colodro-Conde, L., Burt, S. A., Klump, K. L., Medland, S. E., Montgomery, G. W., Kandler, C., McAdams, T. A.,… Kaprio, J. (2015). The CODATwins Project: The Cohort Description of Collaborative Project of Development of Anthropometrical Measures in Twins to Study Macro-Environmental Variation in Genetic and Environmental Effects on Anthropometric Traits. Twin research and human genetics : the official journal of the International Society for Twin Studies, 18(4), 348–360. 10.1017/thg.2015.29

16. Silventoinen, K., Jelenkovic, A., Yokoyama, Y., Sund, R., Sugawara, M., Tanaka, M., Matsumoto, S., Bogl, L. H., Freitas, D. L., Maia, J. A., Hjelmborg, J. V. B., Aaltonen, S., Piirtola, M., Latvala, A., Calais-Ferreira, L., Oliveira, V. C., Ferreira, P. H., Ji, F., Ning, F., Pang, Z.,… Kaprio, J. (2019). The CODATwins Project: The Current Status and Recent Findings of COllaborative Project of Development of Anthropometrical Measures in Twins. Twin research and human genetics : the official journal of the International Society for Twin Studies, 22(6), 800–808. 10.1017/thg.2019.35

17. Halloran E. C. (2024). Adult Development and Associated Health Risks. Journal of patient-centered research and reviews, 11(1), 63–67. 10.17294/2330-0698.2050

18. Medley M. L. (1980). Life satisfaction across four stages of adult life. International journal of aging & human development, 11(3), 193–209. 10.2190/D4LG-ALJQ-8850-GYDV

19. Pinheiro, J., & Bates, D. (2006). Mixed-effects models in S and S-PLUS. Springer science & business media.

20. Posthuma, D., Beem, A. L., de Geus, E. J., van Baal, G. C., von Hjelmborg, J. B., Iachine, I., & Boomsma, D. I. (2003). Theory and practice in quantitative genetics. Twin research : the official journal of the International Society for Twin Studies, 6(5), 361–376. 10.1375/136905203770326367

21. Kaprio, J., & Silventoinen, K. (2011). Advanced methods in twin studies. Genetic Epidemiology, 143–152.

22. Neale, M. C., Hunter, M. D., Pritikin, J. N., Zahery, M., Brick, T. R., Kirkpatrick, R. M.,… & Boker, S. M. (2016). OpenMx 2.0: Extended structural equation and statistical modeling. Psychometrika, 81, 535–549.

23. Li, S., Kyvik, K. O., Pang, Z., Zhang, D., Duan, H., Tan, Q.,… & Dalgård, C. (2016). Genetic and environmental regulation on longitudinal change of metabolic phenotypes in Danish and Chinese adult twins. Plos one, 11(2), e0148396.

24. Fabsitz RR, Sholinsky P, Carmelli D. Genetic influences on adult weight gain and maximum body mass index in male twins. Am J Epidemiol. 1994 Oct 15;140(8):711–20.

25. Austin MA, Friedlander Y, Newman B, Edwards K, Mayer-Davis EJ, King MC. Genetic influences on changes in body mass index: a longitudinal analysis of women twins. Obes Res. 1997;5(4):326–31.

26. Prado, C.M., Batsis, J.A., Donini, L.M. et al. Sarcopenic obesity in older adults: a clinical overview. Nat Rev Endocrinol 20, 261–277 (2024). 10.1038/s41574-023-00943-z

