## Supplementary figure and Tables for "Genetic and environmental effects on weight gain from young adulthood to old age and its association with body mass index at early young adulthood: an individual-based pooled analysis of 16 twin cohorts"

**Supplementary figure 1.** BMI changes distribution for identifying outliers.

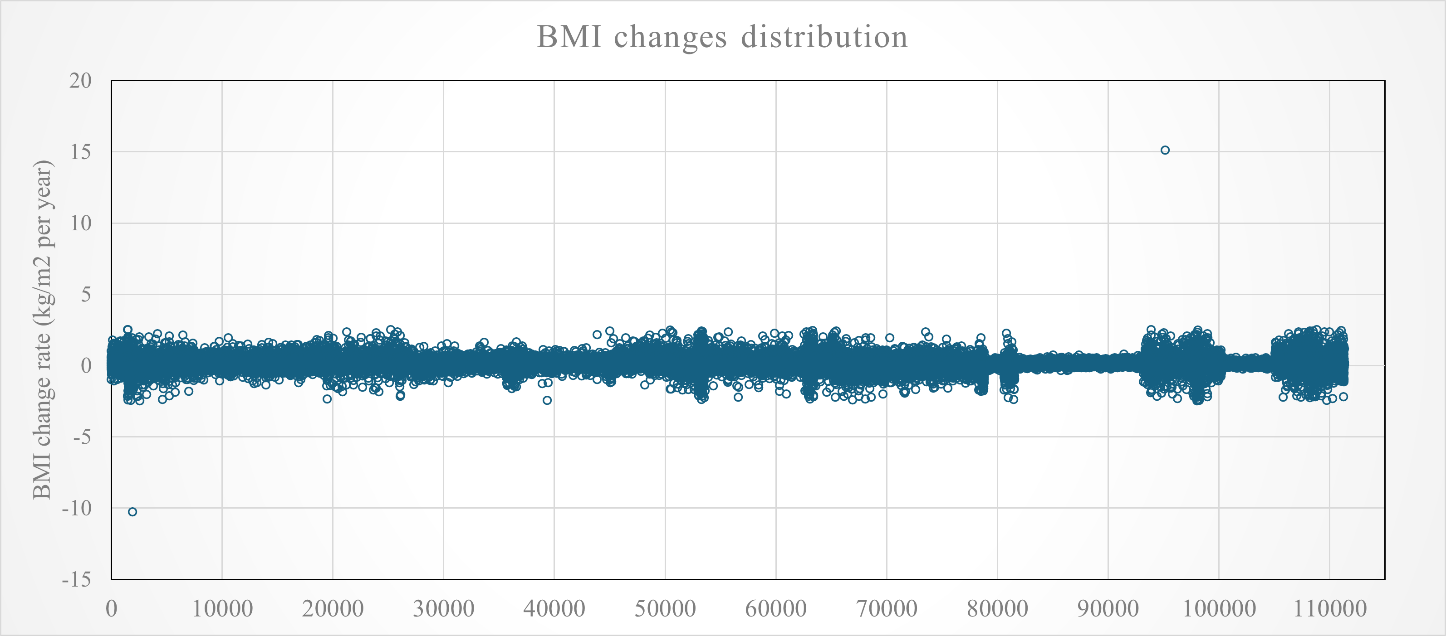

**Caption:** Scatter plot displaying BMI changes values across individuals is shown in the figure. Those individuals that seemed to be outliers in the scatterplot were removed before running the analysis. **Abbreviations:** BMI: body mass index.

**Supplementary table 1.** Descriptive statistics of the cohorts included in the study before applying exclusion criteria.

|  | General information | | | Follow-up | | Zygosity | | | |
| --- | --- | --- | --- | --- | --- | --- | --- | --- | --- |
|  | Age range | N of individuals | % women | N of measures | Mean follow-up time | Complete twin pairs | MZ | DZ | |
|  |  |  |  |  |  |  |  | SSDZ | OSDZ |
| **Europe** |  |  |  |  |  |  |  |  |  |
| Danish Twin Cohort | 18–100 | 28,331 | 54% | 6 | 9.25 | 9,063 | 32% | 41% | 27% |
| Swedish Twin Register | 18–99 | 34,279 | 55% | 4 | 19.90 | 13,860 | 39% | 61% | 0% |
| Finnish old cohort | 18–100 | 21,578 | 52% | 4 | 19.21 | 9,375 | 34% | 66% | 0% |
| Finntwin12 | 18–39 | 1,659 | 60% | 3 | 13.00 | 302 | 61% | 20% | 19% |
| Finntwin16 | 18–37 | 5,003 | 55% | 4 | 11.82 | 2,306 | 33% | 38% | 29% |
| TwinsUK | 18–89 | 6,917 | 91% | 8 | 7.39 | 3,066 | 51% | 47% | 2% |
| Netherlands Twin Cohort | 18–91 | 7,459 | 66% | 13 | 10.65 | 3,338 | 54% | 30% | 16% |
| Italian Twin Registry | 18–78 | 1,433 | 66% | 4 | 4.23 | 455 | 60% | 29% | 11% |
| Murcia Twin Registry | 33–73 | 2,086 | 64% | 3 | 5.51 | 749 | 45% | 36% | 19% |
| Swedish Young Male Twin Study | 18–30 | 2,081 | 0% | 3 | 6.57 | 1,015 | 58% | 42% | 0% |
| **North America** |  |  |  |  |  |  |  |  |  |
| Washington State Twin Registry | 18–97 | 6,588 | 66% | 6 | 6.67 | 3,135 | 59% | 27% | 14% |
| Colorado Twin Registry | 18–34 | 1,683 | 56% | 3 | 6.62 | 746 | 49% | 30% | 21% |
| NAS-NRC Twin Registry | 18–82 | 15,480 | 0% | 4 | 22.60 | 2,400 | 54% | 46% | 0% |
| Vietnam Era Twin Study of Aging | 51–66 | 989 | 0% | 2 | 5.76 | 451 | 60% | 40% | 0% |
| **Asia** |  |  |  |  |  |  |  |  |  |
| Korea Twin-Family Register | 21–79 | 913 | 65% | 4 | 4.37 | 425 | 83% | 17% | 0% |
| **Australia** |  |  |  |  |  |  |  |  |  |
| Queensland Twin Register | 18–92 | 11,859 | 61% | 9 | 14.00 | 4,971 | 44% | 36% | 20% |
| **World** |  |  |  |  |  |  |  |  |  |
| All cohorts | 18–100 | 148,338 | 42% | 2-13 | 15.83 | 55,657 | 42% | 46% | 12% |

**Caption:** Descriptive information of the 16 databases that form the CODATwins database used in the current study after excluding first, those cohorts without information in adult life and, second, individuals with at least two longitudinal measures of BMI. The information displayed in the table is before the application of exclusion criteria**. Abbreviations**: N: number; SD: Standard deviation; MZ: Monozygotic; DZ: Dizygotic; SSDZ: Same sex dizygotic; OSDZ: opposite sex dizygotic; NAS-NRC: National Academy of Sciences-National Research Council.

**Supplementary table 2.** Information on baseline age, baseline BMI and BMI changes in each of the 16 cohorts included in the study by sex.

| Cohorts of study | Men | | | | | | Women | | | | | |
| --- | --- | --- | --- | --- | --- | --- | --- | --- | --- | --- | --- | --- |
|  | Age baseline | | BMI baseline | | BMI changes | | Age baseline | | BMI baseline | | BMI changes | |
|  | Mean | SD | Mean | SD | Mean | SD | Mean | SD | Mean | SD | Mean | SD |
| **Europe** |  |  |  |  |  |  |  |  |  |  |  |  |
| Danish Twin Cohort | 40.26 | 15.91 | 24.45 | 3.12 | 0.15 | 0.26 | 39.51 | 16.71 | 22.77 | 3.61 | 0.15 | 0.29 |
| Swedish Twin Register | 40.25 | 13.08 | 23.73 | 2.77 | 0.05 | 0.22 | 41.03 | 13.36 | 22.86 | 3.49 | 0.06 | 0.23 |
| Finnish Old Cohort | 32.43 | 11.58 | 23.67 | 2.97 | 0.10 | 0.17 | 33.44 | 13.04 | 22.33 | 3.39 | 0.11 | 0.20 |
| FinnTwin12 | 23.06 | 2.29 | 23.83 | 3.10 | 0.19 | 0.22 | 23.20 | 2.18 | 22.40 | 3.76 | 0.20 | 0.25 |
| FinnTwin16 | 18.09 | 1.38 | 21.93 | 2.50 | 0.27 | 0.25 | 18.73 | 1.09 | 20.91 | 2.74 | 0.20 | 0.26 |
| TwinsUK | 48.96 | 14.38 | 25.45 | 3.25 | 0.04 | 0.25 | 47.18 | 11.99 | 24.67 | 4.36 | 0.08 | 0.40 |
| Netherland Twin Cohort | 27.62 | 13.49 | 22.48 | 3.02 | 0.17 | 0.25 | 29.43 | 13.78 | 22.16 | 5.35 | 0.13 | 0.23 |
| Italian Twin Registry | 36.29 | 15.01 | 24.02 | 3.54 | 0.12 | 0.31 | 33.13 | 13.65 | 21.69 | 3.50 | 0.11 | 0.33 |
| Murcia Twin Registry | 50.69 | 5.23 | 27.12 | 3.90 | 0.05 | 0.50 | 48.36 | 5.62 | 25.10 | 4.10 | 0.13 | 0.57 |
| Swedish Young Male Twin Study | 18.41 | 1.15 | 21.63 | 2.54 | 0.31 | 0.37 | - | - | - | - | - | - |
| **North America** |  |  |  |  |  |  |  |  |  |  |  |  |
| Washington State Twin Registry | 40.16 | 18.68 | 25.56 | 3.94 | 0.11 | 0.41 | 37.67 | 17.25 | 24.31 | 4.31 | 0.14 | 0.42 |
| Colorado Twin Registry | 19.67 | 1.64 | 23.18 | 3.28 | 0.26 | 0.34 | 19.74 | 1.63 | 21.93 | 3.45 | 0.22 | 0.41 |
| NAS-NRC Twin Registry | 20.83 | 4.74 | 22.19 | 2.42 | 0.09 | 0.07 | - | - | - | - | - | - |
| Vietnam Era Twin Study of Aging | 55.71 | 2.46 | 28.88 | 4.37 | 0.08 | 0.33 | - | - | - | - | - | - |
| **Asia** |  |  |  |  |  |  |  |  |  |  |  |  |
| Korea Twin-Family Register | 40.04 | 7.93 | 24.23 | 2.58 | 0.03 | 0.22 | 38.91 | 7.38 | 22.34 | 2.89 | 0.06 | 0.27 |
| **Australia** |  |  |  |  |  |  |  |  |  |  |  |  |
| Queensland Twin Register | 28.42 | 10.03 | 23.48 | 2.87 | 0.17 | 0.28 | 30.13 | 11.38 | 22.31 | 3.14 | 0.16 | 0.26 |

**Caption:** Mean value and Standard deviation of baseline age, baseline BMI and BMI changes in each of the 16 cohorts included in the study is displayed in men and women. **Abbreviations:** BMI: Body mass index; SD: Standard deviation; NAS-NRC: National Academy of Sciences-National Research Council.

**Supplementary table 3.** Means and standard deviations of body mass index (BMI) changes (kg/m^2^ per year) across different stages of life in the various databases used by sex.

|  | Men | | Women | | Sex differences (p-value) |
| --- | --- | --- | --- | --- | --- |
|  | Mean | SD | Mean | SD |  |
| **Europe** |  |  |  |  |  |
| *Danish Twin Cohort* |  |  |  |  |  |
| Early young adulthood | 0.26 | 0.39 | 0.18 | 0.43 | 0.04 |
| Late young adulthood | 0.17 | 0.24 | 0.23 | 0.38 | 2.92e-03 |
| Early middle age | 0.05 | 0.40 | 0.05 | 0.48 | 0.16 |
| Late middle age | 0.06 | 0.35 | 0.06 | 0.37 | 0.10 |
| Early old age | -0.02 | 0.59 | -0.10 | 0.72 | 0.02 |
| Late old age | 0.07 | 0.47 | 0.03 | 0.60 | 0.16 |
| *Swedish Twin Register* |  |  |  |  |  |
| Early young adulthood | - | - | - | - | - |
| Late young adulthood | - | - | - | - | - |
| Early middle age | 0.01 | 0.02 | 0.03 | 0.02 | 6.79e-08 |
| Late middle age | 0.01 | 0.02 | 0.02 | 0.02 | 2.74e-15 |
| Early old age | 0.01 | 0.04 | 0.02 | 0.03 | 0.03 |
| Late old age | -0.01 | 0.04 | 6.96e-03 | 0.04 | 0.47 |
| *Finnish Old Cohort* |  |  |  |  |  |
| Early young adulthood | 0.19 | 0.27 | 0.12 | 0.28 | <2.00e-16 |
| Late young adulthood | 0.07 | 0.23 | 0.14 | 0.27 | 1.97e-03 |
| Early middle age | 0.08 | 0.21 | 0.11 | 0.25 | 0.22 |
| Late middle age | 0.02 | 0.20 | 0.13 | 0.25 | 2.59e-04 |
| Early old age | 7.86e-03 | 0.28 | -0.08 | 0.36 | 1.24e-04 |
| Late old age | -0.04 | 0.22 | -0.22 | 0.42 | 0.01 |
| *FinnTwin12* |  |  |  |  |  |
| Early young adulthood | 0.43 | 0.31 | 0.26 | 0.41 | 0.03 |
| Late young adulthood | - | - | - | - | - |
| Early middle age | - | - | - | - | - |
| Late middle age | - | - | - | - | - |
| Early old age | - | - | - | - | - |
| Late old age | - | - | - | - | - |
| *FinnTwin16* |  |  |  |  |  |
| Early young adulthood | 0.29 | 0.38 | 0.29 | 0.40 | <2.00e-16 |
| Late young adulthood | - | - | - | - | - |
| Early middle age | - | - | - | - | - |
| Late middle age | - | - | - | - | - |
| Early old age | - | - | - | - | - |
| Late old age | - | - | - | - | - |
| *TwinsUK* |  |  |  |  |  |
| Early young adulthood | 0.08 | 0.19 | 0.10 | 0.83 | 0.93 |
| Late young adulthood | 0.04 | 0.16 | 0.10 | 0.90 | 0.81 |
| Early middle age | 0.16 | 0.64 | 0.11 | 1.01 | 0.34 |
| Late middle age | 0.02 | 0.13 | 0.07 | 0.40 | 0.27 |
| Early old age | 0.09 | 0.73 | -0.03 | 0.67 | 0.22 |
| Late old age | 0.02 | 8.01e-03 | 0.04 | 0.02 | 0.02 |
| *Netherlands Twin Cohort* |  |  |  |  |  |
| Early young adulthood | 0.13 | 0.66 | 0.13 | 0.87 | 0.50 |
| Late young adulthood | -0.156 | 1.79 | 0.30 | 1.77 | 0.01 |
| Early middle age | -0.41 | 1.61 | 0.21 | 1.56 | 0.01 |
| Late middle age | -0.06 | 1.09 | 0.16 | 1.27 | 0.04 |
| Early old age | 0.10 | 0.29 | 0.12 | 0.50 | 0.03 |
| Late old age | -0.21 | 0.94 | 0.29 | 0.67 | 0.02 |
| *Italian Twin Registry* |  |  |  |  |  |
| Early young adulthood | 0.10 | 0.08 | 0.15 | 0.12 | 0.09 |
| Late young adulthood | 0.27 | 0.89 | 0.21 | 0.82 | 0.58 |
| Early middle age | 0.04 | 0.20 | 0.22 | 0.59 | 2.36e-03 |
| Late middle age | 0.10 | 0.25 | 0.22 | 0.40 | 0.01 |
| Early old age | 0.05 | 0.03 | 0.48 | 0.98 | 4.01e-04 |
| Late old age | - | - | - | - | - |
| *Murcia Twin Registry* |  |  |  |  |  |
| Early young adulthood | - | - | - | - | - |
| Late young adulthood | - | - | - | - | - |
| Early middle age | 0.04 | 1.25 | 0.11 | 0.54 | 1.28e-03 |
| Late middle age | -0.02 | 0.58 | 0.15 | 0.41 | 2.45e-05 |
| Early old age | - | - | - | - | - |
| Late old age | - | - | - | - | - |
| *Swedish Young Male Twin Study* |  |  |  |  |  |
| Early young adulthood | 0.29 | 0.50 | - | - | - |
| Late young adulthood | - | - | - | - | - |
| Early middle age | - | - | - | - | - |
| Late middle age | - | - | - | - | - |
| Early old age | - | - | - | - | - |
| Late old age | - | - | - | - | - |
| **North America** |  |  |  |  |  |
| *Washington State Twin Registry* |  |  |  |  |  |
| Early young adulthood | 0.10 | 0.04 | 0.14 | 0.06 | 4.99e-10 |
| Late young adulthood | 0.08 | 0.02 | 0.13 | 0.05 | 1.45e-08 |
| Early middle age | 0.10 | 0.04 | 0.11 | 0.08 | 1.01e-04 |
| Late middle age | 0.07 | 0.05 | 0.10 | 0.06 | <2.00e-16 |
| Early old age | 0.06 | 0.04 | 0.08 | 0.05 | 5.63e-08 |
| Late old age | 0.03 | 0.04 | 0.07 | 0.04 | 1.18e-03 |
| *Colorado Twin Registry* |  |  |  |  |  |
| Early young adulthood | 0.26 | 0.46 | 0.23 | 0.54 | 0.11 |
| Late young adulthood | - | - | - | - | - |
| Early middle age | - | - | - | - | - |
| Late middle age | - | - | - | - | - |
| Early old age | - | - | - | - | - |
| Late old age | - | - | - | - | - |
| *NAS-NRC Twin Registry* |  |  |  |  |  |
| Early young adulthood | - | - | - | - | - |
| Late young adulthood | - | - | - | - | - |
| Early middle age | - | - | - | - | - |
| Late middle age | 0.02 | 0.16 | - | - | - |
| Early old age | - | - | - | - | - |
| Late old age | - | - | - | - | - |
| *Vietnam Era Twin Study of Aging* |  |  |  |  |  |
| Early young adulthood | - | - | - | - | - |
| Late young adulthood | - | - | - | - | - |
| Early middle age | - | - | - | - | - |
| Late middle age | 0.09 | 0.37 | - | - | - |
| Early old age | - | - | - | - | - |
| Late old age | - | - | - | - | - |
| **Asia** |  |  |  |  |  |
| *Korea Twin-Family Register* |  |  |  |  |  |
| Early young adulthood | - | - | - | - | - |
| Late young adulthood | 0.02 | 0.05 | 0.07 | 0.05 | 8.52e-05 |
| Early middle age | -0.01 | 0.03 | 0.08 | 0.06 | 4.18e-06 |
| Late middle age | 7.75e-03 | 0.04 | 0.06 | 0.05 | 2.90e-05 |
| Early old age | - | - | - | - | - |
| Late old age | - | - | - | - | - |
| **Australia** |  |  |  |  |  |
| *Queensland* |  |  |  |  |  |
| Early young adulthood | 0.21 | 0.31 | 0.15 | 0.38 | 7.47e-06 |
| Late young adulthood | 0.11 | 0.35 | 0.21 | 0.37 | 2.10e-03 |
| Early middle age | 0.08 | 0.16 | 0.14 | 0.26 | 0.03 |
| Late middle age | 0.09 | 0.27 | 0.07 | 0.19 | 0.05 |
| Early old age | -0.54 | 1.42 | -0.63 | 1.45 | 0.35 |
| Late old age | -0.19 | 0.83 | -0.12 | 1.05 | 0.83 |

**Caption:** The body mass index change estimates were obtained by linear regression model based for those with at least three measures and delta method for those with maximum of two measures. The sex difference was studied using linear mixed effect models (model: BMI changes= baseline age +baseline BMI + sex + zygosity + (1|family ID). **Abbreviations**: SD: Standard deviation; BMI: Body mass index: NAS-NRC: National Academy of Sciences-National Research Council.

**Supplementary table 4.** Sensitivity analysis of associations between body mass index (BMI) at early young adulthood and the changes in BMI (kg/m^2^ per year) from early young adulthood to subsequent stage in three phases of life in the pooled data by sex.

| Pairs of Variables to study | Men | | | Women | | |
| --- | --- | --- | --- | --- | --- | --- |
|  | Phenotypic association | | | Phenotypic association | | |
|  | Estimate | 95%CI | | Estimate | 95%CI | |
|  |  | LL | UL |  | LL | UL |
| BMI vs changes early young adulthood-late middle age | -4.65e-03 | -5.36e-03 | -2.17e-06 | 1.71e-03 | 6.45e-05 | 7.04e-03 |
| BMI vs changes early young adulthood-old age | -5.95e-03 | -7.87e-03 | -4.86e-08 | 7.58e-03 | 4.21e-05 | 0.01 |

**Caption**: Associations estimates were obtained from the linear mixed effects model BMI changes in adulthood= BMI at baseline + age at baseline + zygosity + cohort ID + (1|family ID). it is summarized with associations estimates besides their confidence intervals. Abbreviations: CI: Confidence interval; LL: lower limit; UL: upper limit; BMI: body mass index.

**Supplementary table 5.** Sensitivity analysis of associations between body mass index (BMI) at early young adulthood and the changes in BMI (kg/m^2^ per year) from early young adulthood to subsequent stage in six stages of life in the pooled data by sex.

| Pairs of Variables to study | Men | | | Women | | |
| --- | --- | --- | --- | --- | --- | --- |
|  | Phenotypic association | | | Phenotypic association | | |
|  | Estimate | 95%CI | | Estimate | 95%CI | |
|  |  | LL | UL |  | LL | UL |
| BMI vs changes early young adulthood-late young adulthood | -5.67e-03 | -7.85e-03 | -1.27e-06 | 1.30e-03 | -6.24e-04 | 2.23e-03 |
| BMI vs changes early young adulthood- early middle age | 4.78e-03 | 3.45e-05 | 6.04e-03 | 9.43e-03 | 3.04e-08 | 0.01 |
| BMI vs changes early young adulthood-late middle age | -4.65e-03 | -0.01 | -2.45e-05 | 1.71e-03 | 2.26e-05 | 3.86e-03 |
| BMI vs changes early young adulthood-early old age | -6.72e-03 | -8.32e-03 | -4.26e-05 | 7.58e-03 | 1.04e-05 | 0.01 |
| BMI vs changes early young adulthood-late old age | -5.02e-03 | -6.48e-03 | -6.21e-06 | - | - | - |

**Caption**: Associations estimates were obtained from the linear mixed effects model BMI changes in adulthood= BMI at baseline + age at baseline + zygosity + cohort ID + (1|family ID). it is summarized with associations estimates besides their confidence intervals. Abbreviations: CI: Confidence interval; LL: lower limit; UL: upper limit; BMI: body mass index.

**Supplementary table 6.** Intraclass correlations for body mass index (BMI) changes (kg/m^2^ per year) of three stages of adulthood in pooled data by sex and zygosity.

| Variables of study | Men | | | | | | Women | | | | | | Opposite sex dizygotic | | |
| --- | --- | --- | --- | --- | --- | --- | --- | --- | --- | --- | --- | --- | --- | --- | --- |
|  | MZ | | | DZ | | | MZ | | | DZ | | |  |  |  |
|  | Estimate | 95% CI | | Estimate | 95% CI | | Estimate | 95% CI | | Estimate | 95% CI | | Estimate |  | |
|  |  | LL | UL |  | LL | UL |  | LL | UL |  | LL | UL |  | LL | UL |
|  | N of pairs= 1,714 | | | N of pairs= 1,396 | | | N of pairs= 3,435 | | | N of pairs= 2,552 | | | N of pairs= 1,545 | | |
| Young adulthood-early middle age | 0.30 | 0.25 | 0.34 | 0.14 | 0.08 | 0.19 | 0.23 | 0.20 | 0.26 | 0.15 | 0.11 | 0.19 | 0.14 | 0.09 | 0.19 |
|  | N of pairs= 831 | | | N of pairs= 993 | | | N of pairs= 1,014 | | | N of pairs= 1,295 | | | N of pairs= 402 | | |
| Late middle age | 9.56e-04 | -0.06 | 0.06 | 0.11 | 0.05 | 0.17 | 0.14 | 0.08 | 0.20 | 0.06 | 0.01 | 0.12 | 0.01 | -0.04 | 0.23 |
|  | N of pairs= 158 | | | N of pairs= 190 | | | N of pairs= 268 | | | N of pairs= 323 | | | N of pairs= 47 | | |
| Old age | 0.07 | -0.08 | 0.22 | 0.12 | -0.01 | 0.26 | 0.19 | 0.07 | 0.30 | 0.06 | -0.04 | 0.17 | 0.03 | 0.07 | 0.55 |

**Caption**: Intraclass correlations for BMI changes of three consecutive stages of adulthood in pooled data from Europe, Asia, Oceania, and North America are summarized by sex and zygosity with correlation coefficients besides their 95% confidence interval. All the correlations are highly significant (all p<0.005). **Abbreviations:** MZ: Monozygotic; DZ: Dizygotic; CI: Confidence interval; LL: Lower limit; UL: Upper limit.

**Supplementary table 7.** Intraclass correlations for body mass index (BMI) changes (kg/m^2^ per year) of six stages of adulthood in the pooled data by sex and zygosity.

| Variables of study | Men | | | | | | Women | | | | | | Opposite sex dizygotic | | |
| --- | --- | --- | --- | --- | --- | --- | --- | --- | --- | --- | --- | --- | --- | --- | --- |
|  | MZ | | | DZ | | | MZ | | | DZ | | |  |  |  |
|  | Estimate | 95% CI | | Estimate | 95% CI | | Estimate | 95% CI | | Estimate | 95% CI | | Estimate | 95% CI | |
|  |  | LL | UL |  | LL | UL |  | LL | UL |  | LL | UL |  | LL | UL |
|  | N of pairs= 1,332 | | | N of pairs= 976 | | | N of pairs= 1,458 | | | N of pairs= 834 | | | N of pairs= 975 | | |
| Early young adulthood | 0.31 | 0.26 | 0.35 | 0.05 | -0.00 | 0.12 | 0.23 | 0.18 | 0.28 | 0.27 | 0.20 | 0.33 | 0.15 | 0.09 | 0.21 |
|  | N of pairs= 155 | | | N of pairs= 136 | | | N of pairs= 364 | | | N of pairs= 232 | | | N of pairs= 139 | | |
| Late young adulthood | 0.07 | -0.08 | 0.22 | -0.01 | -0.18 | 0.15 | 0.30 | 0.21 | 0.39 | 0.23 | 0.11 | 0.35 | 0.06 | -0.10 | 0.22 |
|  | N of pairs= 227 | | | N of pairs= 285 | | | N of pairs= 406 | | | N of pairs= 430 | | | N of pairs= 94 | | |
| Early middle age | 0.06 | -0.06 | 0.18 | 0.14 | 0.02 | 0.25 | -0.02 | -0.12 | 0.06 | -0.01 | -0.11 | 0.08 | 0.12 | -0.07 | 0.32 |
|  | N of pairs= 831 | | | N of pairs= 993 | | | N of pairs= 1,014 | | | N of pairs= 1,295 | | | N of pairs= 402 | | |
| Late middle age | 9.56e-04 | -0.06 | 0.06 | 0.11 | 0.05 | 0.17 | 0.14 | 0.08 | 0.20 | 0.06 | 0.01 | 0.12 | 0.14 | 0.04 | 0.23 |
|  | N of pairs= 108 | | | N of pairs= 148 | | | N of pairs= 187 | | | N of pairs= 231 | | | N of pairs= 38 | | |
| Early old age | 0.03 | -0.15 | 0.22 | 0.12 | -0.04 | 0.27 | 0.32 | 0.18 | 0.44 | 0.14 | 0.02 | 0.27 | 0.25 | -0.06 | 0.53 |
|  | N of pairs= 49 | | | N of pairs= 42 | | | N of pairs= 69 | | | N of pairs= 82 | | | N of pairs= 9 | | |
| Late old age | 0.16 | -0.12 | 0.42 | 0.18 | -0.12 | 0.46 | -0.09 | -0.32 | 0.14 | -0.02 | -0.23 | 0.19 | 0.09 | -0.60 | 0.71 |

**Caption**: Intraclass correlations for BMI changes at six stages of adulthood in pooled data from Europe, Asia, Oceania, and North America are summarized by sex and zygosity with correlation coefficients besides their 95% confidence interval. All the correlations are highly significant (all p<0.005). **Abbreviations:** MZ: Monozygotic; DZ: Dizygotic; CI: Confidence interval; LL: Lower limit; UL: Upper limit.

**Supplementary table 8.** Model fit statistics of body mass index (BMI) changes (kg/m^2^ per year) of three stages of adulthood in the pooled data comparing different genetic models.

| Variables of study | Full ACE model (reference model) | | ACE model without sex-specific genetic effect (1) | | ACE model with same parameter estimates for men and women (2) | | Full AE model (3) | | AE model without sex-specific genetic effect (4) | | AE model with same parameter estimates for men and women (5) | |
| --- | --- | --- | --- | --- | --- | --- | --- | --- | --- | --- | --- | --- |
|  | -2LL | d.f | Δ -2 LL | p value | Δ -2 LL | p value | Δ -2 LL | p value | Δ -2 LL | p value | Δ -2 LL | p value |
| Young adulthood-early middle age | 20058.86 | 18170 | -8.00e-11 | 1 | 218.34 | 4.56e-47 | 0.64 | 0.72 | -3.63e-12 | 1 | 218.25 | 4.04e-48 |
| Late middle age | 6056.95 | 8254 | -1.45e-11 | 1 | 29.68 | 1.60e-06 | 4.14 | 0.12 | -3.63e-12 | 1 | 28.39 | 6.81e-07 |
| Old age | 2406.29 | 1858 | -1.81e-11 | 1 | 8.18 | 0.04 | 0.64 | 0.72 | -4.54e-13 | 1 | 7.53 | 0.02 |

**Caption:** Full ACE model is compared against the rest of the models displaying the -2 log likelihood and degrees of freedom for the reference model and the differences in -2log likelihood and the p value in the models compared with the reference one. **Abbreviations:** -2LL (-2 log-likelihood); d.f. (degrees of freedom); Δ (change); ACE (additive genetic/ shared environment/ unique environment) model; AE (additive genetic/ unique environment) model. (1) Compared to full ACE model (Δ d.f. 1); (2) Compared to ACE model without sex-specific genetic effect (Δ d.f. 2); (3) Compared to full ACE model (Δ d.f. 2); (4) Compared to the full AE model (Δ d.f. 1); (5) Compared to the AE model without sex-specific genetic effect (Δ d.f. 1).

**Supplementary table 9.** Model fit statistics of body mass index (BMI) changes (kg/m^2^ per year) of six stages of adulthood in the pooled data comparing different genetic models.

| Variables of study | Full ACE model (reference model) | | ACE model without sex-specific genetic effect (1) | | ACE model with same parameter estimates for men and women (2) | | Full AE model (3) | | AE model without sex-specific genetic effect (4) | | AE model with same parameter estimates for men and women (5) | |
| --- | --- | --- | --- | --- | --- | --- | --- | --- | --- | --- | --- | --- |
|  | -2LL | d.f | Δ -2 LL | p value | Δ -2 LL | p value | Δ -2 LL | p value | Δ -2 LL | p value | Δ -2 LL | p value |
| Early young adulthood | 9270.02 | 9192 | -1.09e-11 | 1 | 88.28 | 5.11e-19 | 13.53 | 1.15e-04 | -1.81e-12 | 1 | 76.95 | 1.94e-17 |
| Late young adulthood | 1837.33 | 1766 | -6.13e-12 | 1 | 106.21 | 7.16e-23 | 0.33 | 0.84 | -6.82e-13 | 1 | 106.93 | 6.02e-24 |
| Early middle age | 3419.00 | 2679 | -7.91e-11 | 1 | 155.80 | 1.47e-33 | 1.79 | 0.40 | -9.77e-11 | 1 | 156.76 | 1.47e-34 |
| Late middle age | 6056.95 | 8254 | -1.45e-11 | 1 | 29.68 | 1.60e-06 | 4.14 | 0.12 | -3.63e-12 | 1 | 28.39 | 6.81e-07 |
| Early old age | 1418.39 | 1328 | -1.94e-10 | 1 | 5.90 | 0.11 | 0.63 | 0.72 | -2.27e-13 | 1 | 5.28 | 0.07 |
| Late old age | 751.26 | 476 | -7.41e-11 | 1 | 3.18 | 0.36 | -7.79e-11 | 1 | -1.61e-11 | 1 | 3.21 | 0.20 |

**Caption:** Full ACE model is compared against the rest of the models displaying the -2 log likelihood and degrees of freedom for the reference model and the differences in -2log likelihood and the p value in the models compared with the reference one. **Abbreviations:** -2LL (-2 log-likelihood); d.f. (degrees of freedom); Δ (change); ACE (additive genetic/ shared environment/ unique environment) model; AE (additive genetic/ unique environment) model. (1) Compared to full ACE model (Δ d.f. 1); (2) Compared to ACE model without sex-specific genetic effect (Δ d.f. 2); (3) Compared to full ACE model (Δ d.f. 2); (4) Compared to the full AE model (Δ d.f. 1); (5) Compared to the AE model without sex-specific genetic effect (Δ d.f. 1).

**Supplementary table 10**. BMI change rates across six different stages of adulthood with 95% confidence intervals by sex.

| Life stages | Men | | | Women | | |
| --- | --- | --- | --- | --- | --- | --- |
|  | Mean | 95% CI | | Mean | 95%CI | |
|  |  | LL | UL |  | LL | UL |
| Early young adulthood | 0.26 | 0.19 | 0.30 | 0.18 | 0.14 | 0.21 |
| Late young adulthood | 0.13 | 0.07 | 0.18 | 0.14 | 0.07 | 0.19 |
| Early middle age | 0.03 | 0.00 | 0.06 | 0.08 | 0.03 | 0.12 |
| Late middle age | 0.04 | 0.01 | 0.06 | 0.06 | 0.02 | 0.10 |
| Early old age | 0.01 | -0.02 | 0.16 | -0.02 | -0.06 | 0.01 |
| Late old age | -0.06 | -0.11 | -0.01 | -0.01 | -0.04 | 0.02 |

**Caption:** BMI change rate is displayed in early young adulthood, late young adulthood, early middle age, late middle age, early old age and late old age in men and women from CODATwins data before applying exclusion criteria. In both the change rate decrease as one progresses through the phases. **Abbreviations:** CI: Confidence Interval; LL: Lower limit; UL: Upper limit.

**Supplementary table 11**. Relative proportions of body mass index (BMI) changes (kg/m^2^ per year) rate variance explained by additive genetic and unique environmental variance components with 95% confidence intervals of BMI changes among across 6 different stages of adulthood in the pooled data by sex.

|  | Men | | | | | | Women | | | | | |
| --- | --- | --- | --- | --- | --- | --- | --- | --- | --- | --- | --- | --- |
|  | Additive genetic effect | | | Unique environmental effect | | | Additive genetic effect | | | Unique environmental effect | | |
|  | a^2^ | 95%CI | | e^2^ | 95%CI | | a^2^ | 95%CI | | e^2^ | 95%CI | |
|  |  | LL | UL |  | LL | UL |  | LL | UL |  | LL | UL |
| Early young adulthood | 0.28 | 0.23 | 0.32 | 0.72 | 0.67 | 0.76 | 0.29 | 0.23 | 0.32 | 0.71 | 0.67 | 0.76 |
| Late young adulthood | 0.05 | 0.00 | 0.17 | 0.96 | 0.82 | 1.00 | 0.36 | 0.26 | 0.44 | 0.64 | 0.56 | 0.73 |
| Early middle age | 0.12 | 0.01 | 0.22 | 0.88 | 0.77 | 0.98 | 0.09 | 0.00 | 0.18 | 0.91 | 0.81 | 0.99 |
| Late middle age | 0.05 | 0.00 | 0.11 | 0.95 | 0.89 | 1.00 | 0.16 | 0.09 | 0.21 | 0.84 | 0.78 | 0.90 |
| Early old age | 0.09 | 0.00 | 0.25 | 0.91 | 0.74 | 1.00 | 0.33 | 0.20 | 0.44 | 0.67 | 0.55 | 0.79 |
| Late old age | 0.26 | 0.00 | 0.54 | 0.74 | 0.45 | 1.00 | - | - | - | - | - | - |

**Caption:** AE model without sex specific genetic effect was selected for BMI changes in all the different stages of adulthood. The results show an additive genetic component in all stages with women having higher additive genetic components in all stages. **Abbreviations:** a^2^: proportion of total variance explained by additive genetic factors (Heritability); e^2^: proportion of total variance explained by unique environmental factors; CI: Confidence interval; LL: Lower limit; UL: Upper limit.

**Supplementary table 12.** Phenotypic, additive genetic and unique environmental correlations of body mass index (BMI) at early young adulthood and the changes in BMI (kg/m^2^ per year) from early young adulthood to subsequent stage, considering six different stages adulthood in the pooled data sex.

| Pairs of Variables to study | Phenotypic correlation | | | Additive genetic correlation | | | Unique environmental correlation | | |
| --- | --- | --- | --- | --- | --- | --- | --- | --- | --- |
|  | r | 95%CI | | r_A_ |  | | r_E_ |  | |
|  |  | LL | UL |  | LL | UL |  | LL | UL |
| **Men** |  |  |  |  |  |  |  |  |  |
| BMI at early adulthood vs changes early adulthood-late young adulthood | -0.09 | -0.11 | -0.06 | 0.01 | -0.08 | 0.08 | -0.13 | -0.20 | -0.06 |
| BMI at early adulthood vs changes early adulthood-early middle age | 0.06 | 0.02 | 0.10 | 0.24 | -0.01 | 0.43 | -0.11 | -0.22 | -0.01 |
| BMI at early adulthood vs changes early adulthood-late middle age | -0.13 | -0.15 | -0.10 | -0.04 | -0.09 | 0.03 | -0.14 | -0.20 | -0.07 |
| BMI at early adulthood vs changes early adulthood-early old age | -0.21 | -0.25 | -0.18 | -0.26 | -0.35 | -0.26 | -0.21 | -0.28 | -0.14 |
| BMI at early adulthood vs changes early adulthood-late old age | -0.23 | -0.28 | -0.19 | -0.30 | -0.40 | -0.19 | -0.22 | -0.32 | -0.12 |
| **Women** |  |  |  |  |  |  |  |  |  |
| BMI at early adulthood vs changes early adulthood-late young adulthood | 0.02 | 0.00 | 0.05 | 0.35 | 0.20 | 0.47 | -0.09 | -0.13 | -0.04 |
| BMI at early adulthood vs changes early adulthood-early middle age | 0.27 | 0.24 | 0.30 | 0.64 | 0.47 | 0.86 | 0.04 | -0.04 | 0.12 |
| BMI at early adulthood vs changes early adulthood-late middle age | 0.05 | 0.03 | 0.08 | 0.27 | 0.14 | 0.40 | -0.03 | -0.09 | 0.01 |
| BMI at early adulthood vs changes early adulthood-early old age | 0.28 | 0.21 | 0.34 | 0.51 | 0.34 | 0.67 | 0.03 | -0.13 | 0.20 |
| BMI at early adulthood vs changes early adulthood-late old age | - | - | - | - | - | - | - | - | - |

**Caption**: Pairwise correlations between body mass index (BMI) at early young adulthood and the changes in BMI (kg/m^2^ per year) from early young adulthood to subsequent stage, considering six different stages of adulthood were carried out. They are summarized with correlations estimates besides their confidence intervals. Abbreviations: r_A_: additive genetic correlation; r_E_; specific environmental correlation; CI: Confidence interval; LL: lower limit; UL: upper limit; BMI: body mass index.
